# Improving Computational Fluid Dynamics Simulations of Coiled Aneurysms Using Finite Element Modeling

**DOI:** 10.1101/2023.02.27.23286512

**Authors:** Patrick Fillingham, Julia Romero Bhathal, Laurel M.M. Marsh, Michael C. Barbour, Mehmet Kurt, Ciprian N. Ionita, Jason M. Davies, Alberto Aliseda, Michael R. Levitt

## Abstract

Cerebral aneurysms are a serious clinical challenge, with ∼half resulting in death or disability. Treatment via endovascular coiling significantly reduces the chances of rupture, but the technique has failure rates between 25-40%. This presents a pressing need to develop a method for determining optimal coil deployment strategies. Quantification of aneurysm hemodynamics through computational fluid dynamics (CFD) has the potential to significantly improve the understanding of the mechanics of aneurysm coiling and improve treatment outcomes, but accurately representing the coil mass in CFD simulations remains a challenge. We have used the Finite Element Method (FEM) for simulating patient-specific coil deployment based on mechanical properties and coil geometries provided by the device manufacturer for n=4 ICA aneurysms for which 3D printed *in vitro* models were also generated, coiled, and scanned using ultra-high resolution synchrotron micro-CT. The physical and virtual coil geometries were voxelized onto a binary structured grid and porosity maps were generated for geometric comparison. The average binary accuracy score is 0.836 and the average error in porosity map is 6.3%. We then conduct patient-specific CFD simulations of the aneurysm hemodynamics using virtual coils geometries, micro*-*CT generated oil geometries, and using the porous medium method to represent the coil mass. Hemodynamic parameters of interest including were calculated for each of the CFD simulations. The average error across hemodynamic parameters of interest is ∼19%, a 58% reduction from the average error of the porous media simulations, demonstrating a marked improvement in the accuracy of CFD simulations using FEM generated coil geometries.

## INTRODUCTION

Endovascular embolization is becoming a preferred method of treatment for intracranial aneurysms at risk of rupture. Despite growing prevalence of the procedure, predicting and evaluating the efficacy of endovascular treatment remains difficult, while treatment failure rates remain as high as 30%.(1, 2) The primary method of endovascular aneurysm treatment is coil embolization, where the aneurysmal sac is filled with a set of metal coils through an endovascular catheter. The coils reduce flow into the aneurysm, which when coupled with the thrombogenic properties of the coils allows for clotting to occur, leading to a stable thrombus and reduced risk of aneurysm rupture. Whether this flow reduction and thrombosis occur is dependent on the localized hemodynamics of flow in and around the aneurysm that are difficult to predict and impossible to measure *in vivo*.(3, 4, 5)

Computational fluid dynamics (CFD) is a useful tool for analyzing aneurysm hemodynamics that has been instrumental in improving the understanding of aneurysm hemodynamics.(6, 7, 8) However, due to the complex nature of the geometry of coiled aneurysms and the difficulty of capturing this geometry *in vivo*, (9) CFD simulation of coiled aneurysm generally relies on assumptions, such as treating the coiled region as a homogeneous porous medium.(10, 11, 12, 13) While the porous medium models are useful, they cannot capture critical aspects of local hemodynamics that may be significant in treatment failure.(14) Finite element modeling (FEM) can be used to computationally simulate aneurysm coil deployment by solving the equations of motion as the coil is deployed from the catheter into the aneurysm. Recent work (15, 16, 17) has demonstrated the feasibility of simulating virtual coil development using FEM, demonstrating promising qualitative comparisons with coils deployed both *in vivo* and *in vitro*. Due to the difficulty of obtaining highly resolved 3D geometry of physically deployed coils, however, virtual coiling methods have yet to be quantitatively validated in three dimensions and the accuracy of CFD simulations using virtual coils geometries has yet to be investigated.

Our group has previously developed a method for developing anatomically accurate 3D *in vitro* models of coiled cerebral aneurysms (18) which have been scanned using ultra-high-resolution synchrotron microtomography, yielding precise 3D imaging of coil geometry that can be incorporated into CFD simulations.(14) In this work we use FEM to simulate the coiling of patient-specific aneurysm geometries, incorporate them into CFD simulations, and compare the results to CFD simulations using coiled physical models that have been scanned using synchrotron microtomography, as well as simulations representing the coils as a homogeneous porous medium. This permits us to examine the differences in CFD simulations using physical and virtual coil geometries, in an effort to improve their accuracy.

## METHODS

The objective of this work is to investigate the effectiveness of FEM simulation for obtaining accurate deployed coil geometries in coiled cerebral aneurysms, compared to both *in vitro* coil geometries obtained via synchrotron microtomography of 3D printed models, and a standard simplified porous media approximation of the coil mass.

### Subject Selection

Patient-specific aneurysm geometries from n=4 patients treated with embolic coils were selected for this study, which was approved by the institutional review board. Details of the selected aneurysms are summarized in **Table 1**.

**Table 1.**
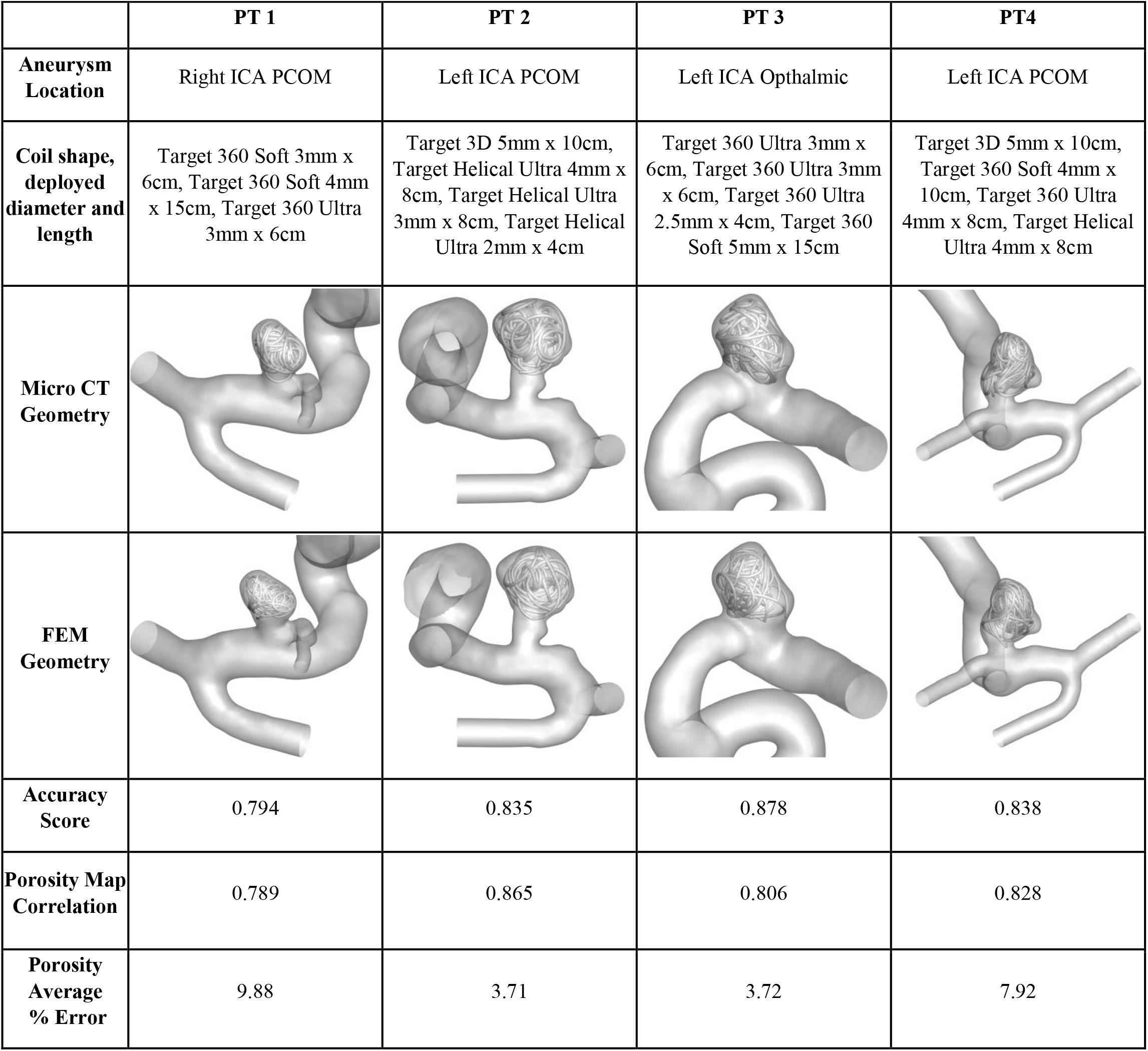
Comparison of FEM and microtomography coil geometries for each of the four patient-specific cerebral aneurysms. Binary voxelization of the geometries are compared via binary accuracy score. Continuous porosity maps are compared via Pearson Correlation coefficient and average error. FEM geometries are align with microtomographic coil geometries with average binary accuracy score of 0.836, average porosity correlation of 0.822 and average error in porosity of 6.3%.

### Finite Element Methodology

#### Patient-Specific Aneurysm Geometry

Patient-specific anatomic information was generated from 3D rotational angiography, and was used to develop both the 3D surface mesh for FEM simulation and the 3D printed i*n vitro* models (18). The *in vitro* models were treated using the same coils (Stryker Neurovascular) as were used in the patient’s clinical care, including temporary balloon remodeling, as previously described (14). As such, a virtual balloon surface matching the curvature of the parent vessel was generated at the neck of each aneurysm geometry. Finally, a virtual catheter geometry was generated and placed at the same location and angle as during the *in vivo* deployments, determined from 3D rotational angiography conducted during deployment. This step is critical in accurately simulating coil deployment, as final coil geometry is highly sensitive to catheter placement.(19) This final geometry is then imported into commercial FEM software (ABAQUS, Dassault Systems) as a discrete rigid geometry.

#### Coil Geometry and Mechanical Properties

In order to accurately simulate coil deployment, it is critical to accurately represent the geometry and mechanical properties of the coils. Endovascular coils contain a multiscale structure, which must be accounted for when simulating coil deployment. The coils consist of a primary wire of diameter *D*_1_ wound tightly into a long helical spring of diameter *D*_2_. The wire-like spring is then wound around a mandrel and heat treated to obtain a desirable 3D structural form or “pre-shape,” with a characteristic diameter *D*3, **(Figure 1)** prior to being pulled into a catheter for packaging and deployment. To limit assumptions, we obtained precise 3D geometries and material properties of all the coils used in this work from the device manufacturer.

**Figure 1.**
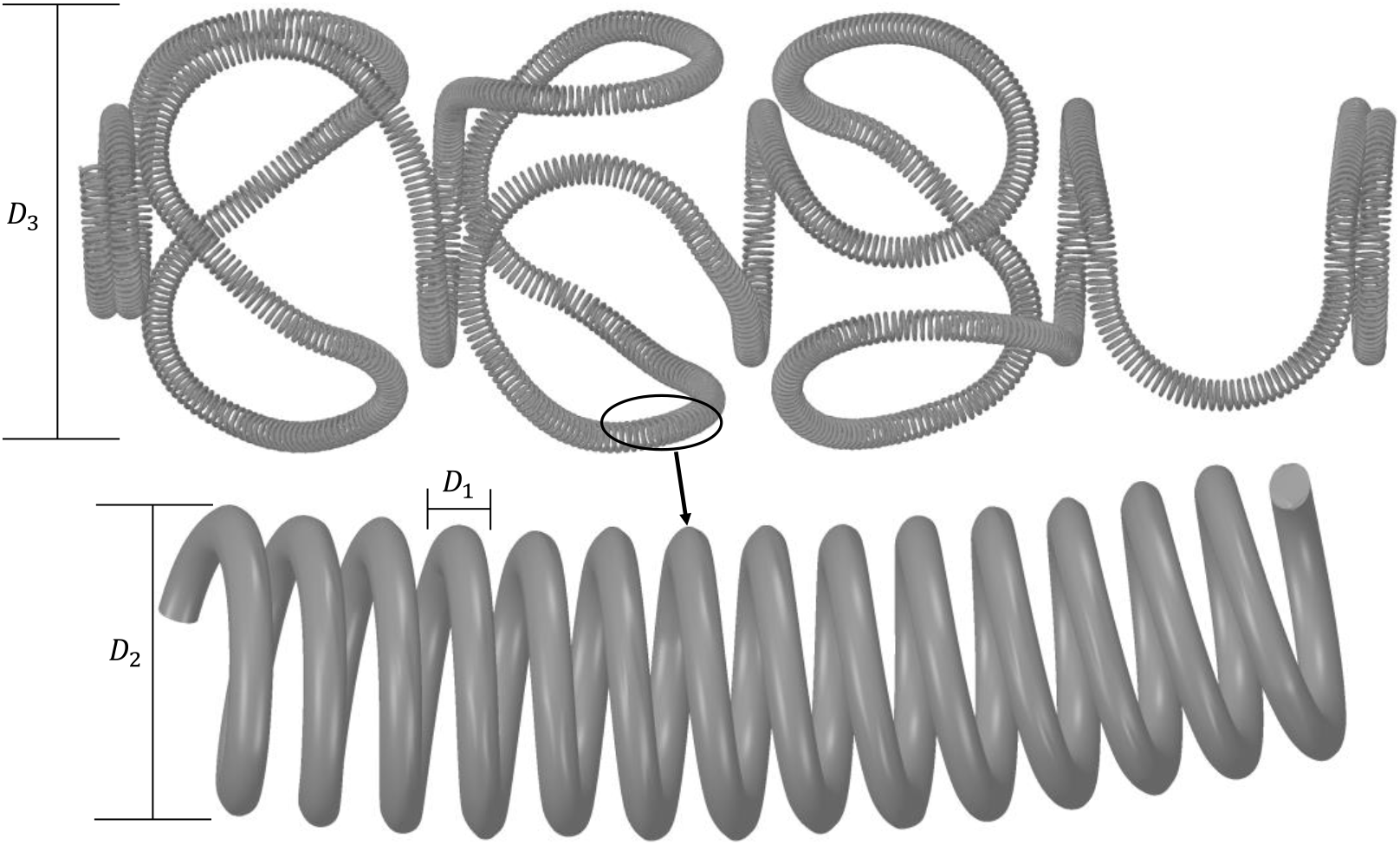
Geometry of a typical endovascular coil preshape (above) with cutout of helical structure of the primary coil (below).

As the resolution necessary to directly model the helical structure of the primary wire of each coil is prohibitive, we instead chose to model the coils as solid beam elements with a diameter of *D*_2_.(16) In order to account for this approximation, the effective Young’s Modulus and Shear Modulus of the beam cross section must be calculated, as using the moduli for the base material would drastically overestimate the stiffness of the coils. In order to calculate these effective moduli we use the formulations first developed by Wahl (20) and successfully implemented for simulating coil deployment by Otani (16) and Damiano (15).

#### Finite Element Modeling Simulation

For each case, the geometry developed for the specific coils used to treat that specific aneurysm (same coil configuration for i*n vivo* and i*n vitro deployment)* was uploaded into the FEM software as discrete deformable wire geometries. Each coil geometry was meshed into beam elements, with a diameter of *D*_2_ and an element length (*dl*) determined by the curvature of the pre-shape such that *dl* = *D*_3_/12, ensuring a minimum of 36 elements per circle, ensuring the unforced angle between beam elements is no greater than 10° while the beam length is still at least 10 times the beam diameter.(21) We used shear-flexible Timoshenko beam elements as they are more appropriate for handling large deformations.(16, 22)

Contact is modeled using the built-in general contact algorithm, which assumes hard contact in the normal direction and a “penalty” method for the tangential contact. Coil-coil contact was modeled with a friction factor of 0.2, commensurate with platinum-platinum contact, and a friction factor of 0.6 for coil-aneurysm and coil-catheter contacts, which is based upon platinum-silicone contact experiments.(19)

The Abaqus/Explicit solver was used to solve the equations of motion during deployment, as it is designed for highly dynamic and nonlinear behavior and uses a lumped mass matrix and a forward Euler central difference algorithm.(21) The simulation is broken down into two dynamic steps for each coil being deployed: First, a packaging step where a dynamic displacement boundary condition, equal in magnitude to the length of the catheter, is applied the distal end of the coil, drawing the coil into the catheter. Second, a deployment step, in which a displacement boundary condition equal to the length of coil is applied to the proximal end of the coil. The packaging step is performed over two virtual seconds, while the deployment step is conducted over 10 seconds, to match the approximate speed of deployment for the *in vitro* deployment. A final dynamic step of 10 seconds is used to remove the catheter from the dome and allow the coil to come to rest. The final coil geometry is then exported as a high-resolution volumetric file for postprocessing.

### Computational Fluid Dynamics Methodology

Our group has extensive experience simulating blood flow through cerebral aneurysms(8, 14, 23, 24) and has previously developed a thorough methodology for conducting CFD simulations incorporating microtomography, and simulations where the coil mass is represented as a homogeneous porous medium.(14) A commercial finite volume solver (Ansys Fluent) was used to solve the transient 3-D Navier-Stokes equations. Blood was assumed to be an incompressible Newtonian fluid with a density of 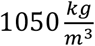 and a viscosity of 3.5 *cP*. Patient specific anatomy was manually segmented from 3D rotational angiography. Patient-specific inlet boundary conditions are derived from *in vivo* dual-sensor doppler guidewires measurements.(8, 25) The coupled velocity and pressure measurements from the guidewire are used to derive a fully resolved Womersley velocity profile at the inlet. Outlet boundary conditions are calculated using a two-element Windkessel model when multiple outlets are present, and a zero-pressure condition when only one outlet is required. Each simulation was conducted with a time step of 0.001 seconds and run for a minimum of five cardiac cycles to eliminate the effect of initial conditions. Simulations were run three times for each patient using the same anatomy and boundary conditions, using three different representations of the coil geometry: The FEM-generated coil geometry, the coil geometry obtained via synchrotron microtomography of 3D printed *in vitro* aneurysm models,(18) and a homogeneous porous medium, as previously described.(14)

## RESULTS

### Comparison of Coil Geometries

Coil geometries were successfully generated for each of the subjects in each condition. Comparisons of the FEM generated and *in vitro* microtomography derived coil geometries are shown in **Figure 1**. To quantitatively analyze the similarity between the coil geometries, a structured 3D grid was generated for each aneurysm, with 100 grid points spanning the diagonal of the bounding box for each aneurysm (yielding an average grid size ∼0.05mm or approximately 1/5^th^ of *D*_2_), onto which both the FEM and microtomography geometries were voxelized, generating a binary 3D matrix for each coil geometry. We then generated an accuracy score for each binary matrix (percentage of binary voxels that match between the two geometries), shown in **Table 1**. As local porosity has been demonstrated to dictate coiled aneurysm hemodynamics (26) we generated porosity maps for each of the coil geometries by calculating the porosity in every 5×5×5 cube in the binary voxelized grid such that the face of each cube is approximately the size of the coil cross section. We then calculated the Pearson’s Correlation Coefficient and average error for each of the porosity maps, also shown in **Table 1**.

### CFD Results

CFD simulations were successfully completed in all three coil modeling conditions for all four patients. While there are quantitative differences between CFD using FEM and microtomography coil geometries, the qualitative similarity is notable. Time averaged wall shear stress contours are plotted side by side in **Figure 2** demonstrate the overall similarity of the results using the FEM and microtomography coil geometries.

**Figure 2.**
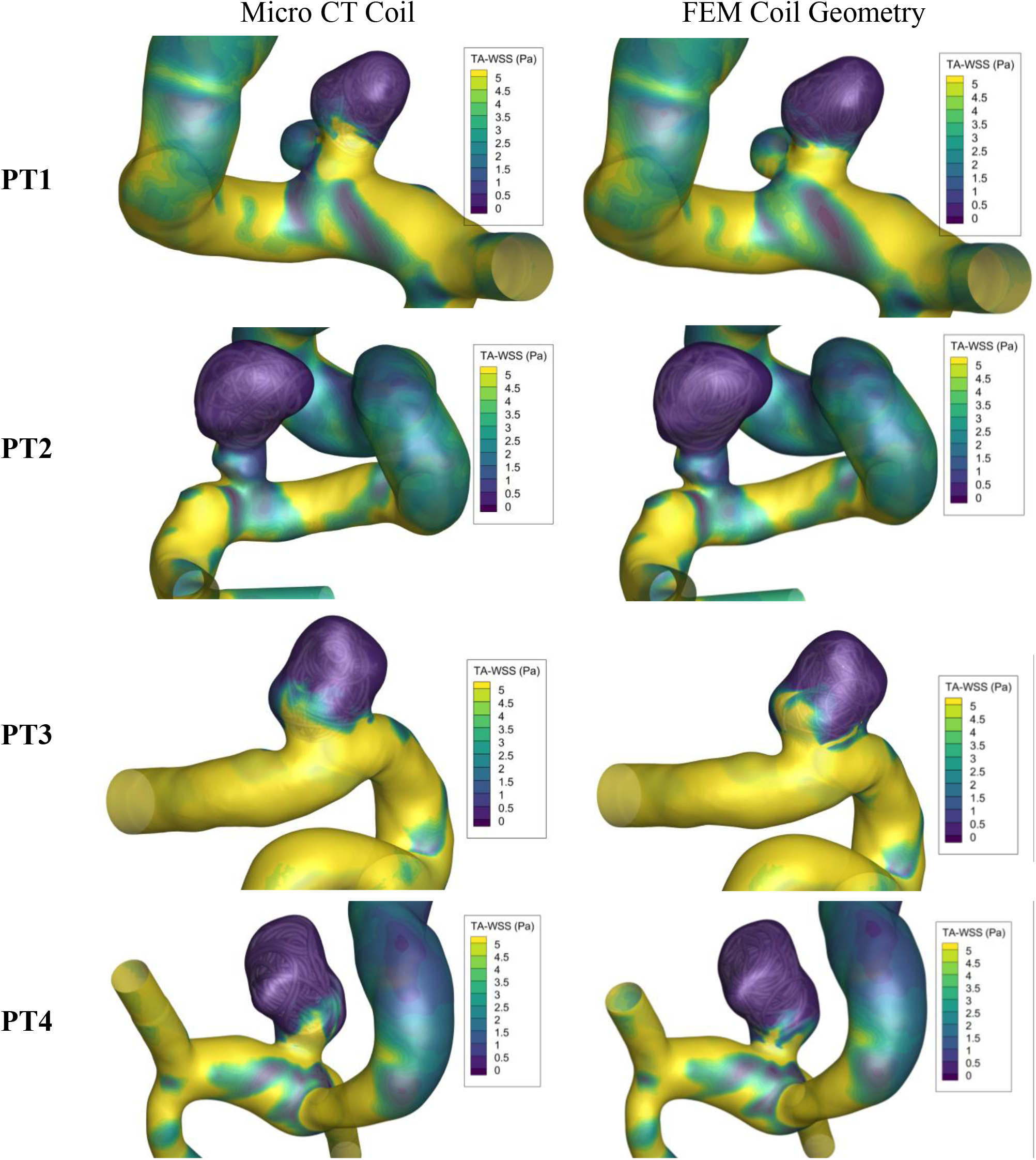
Side-by-side comparison of time-averaged wall shear stress (TA-WSS) using microtomography derived (left) and FEM generated (right) coil geometries in each patient. The spatial distribution and magnitude are generally similar, while there are minor differences in the shear stress maxima caused by the precise orientation of the coil mass.

After CFD simulations were completed, the following hemodynamic parameters of interest (27) were calculated for each of the three simulation conditions conducted for each subject: The average velocity magnitude at the neck of the aneurysm 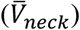, the average velocity magnitude in the aneurysm dome 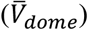, the average wall shear stress across the aneurysm dome 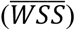, the average of the spatial gradient of wall shear stress across the aneurysm dome 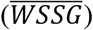, and the oscillatory shear index (*OSI*). With the exception of OSI which is inherently a time averaged value, each of the hemodynamic parameters was calculated as both time average across the entire cardiac cycle, and separately at peak systole. The results of the CFD simulations are shown in **Table 2**. CFD using the FEM coil geometries consistently outperforms the porous media simulations by an average error reduction of ∼58%

**Table 2.**
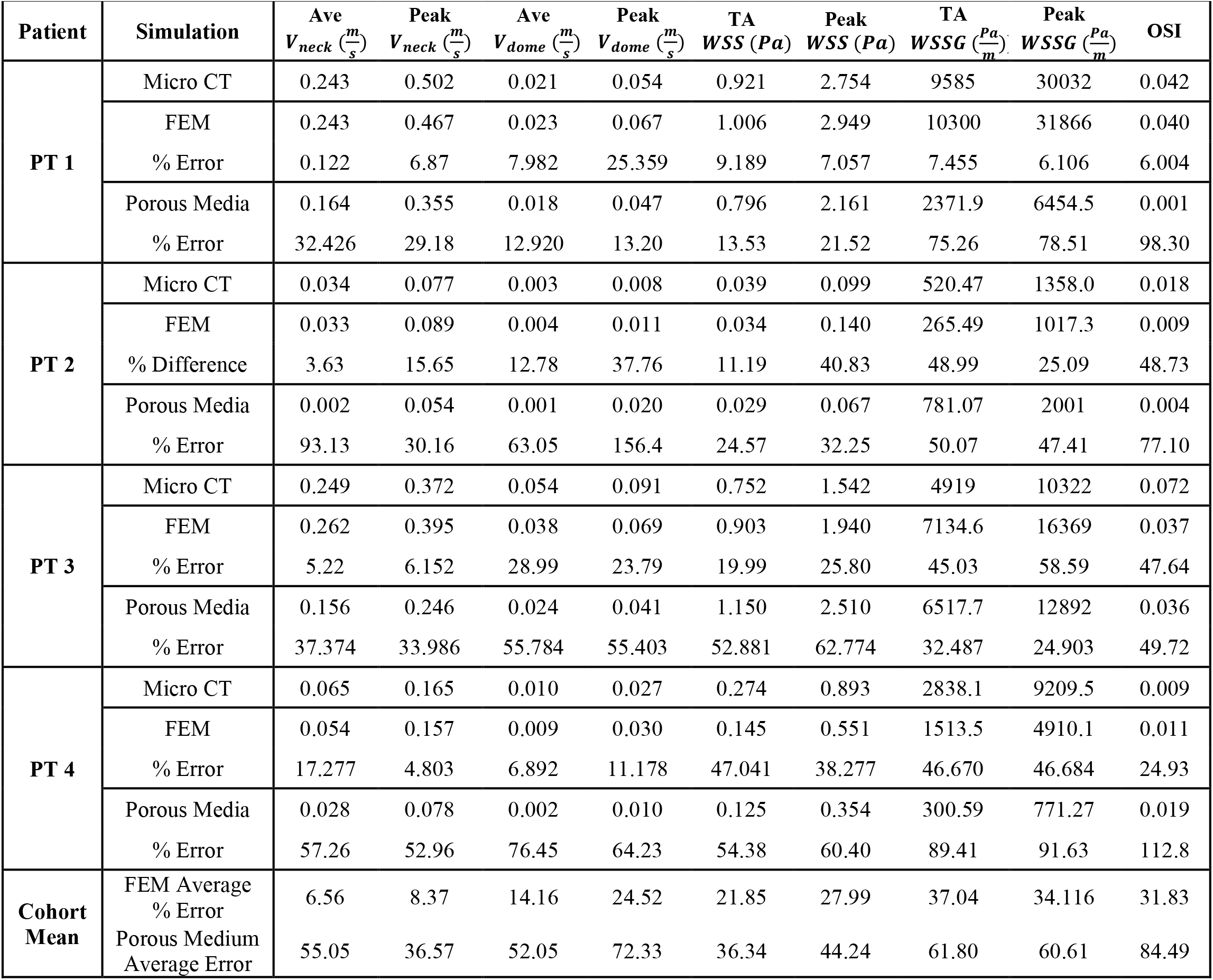
Comparison of hemodynamic parameters of interest derived from CFD using coil geometries defined by microtomography, FEM simulations or porous medium representations. Compared to the microtomographic ground truth coil geometries, the FEM coil geometries significantly outperform the porous media method, with an average error reduction of ∼58%. Ave = time-averaged; peak = peak systole; WSS = wall shear stress; WSSG = wall shear stress gradient; OSI = oscillatory shear index; Micro CT = microtomography coil geometry; FEM = FEM coil geometry

## DISCUSSION

A variety of studies have establish a link between elevated WSS, OSI and velocity at the aneurysm neck and aneurysm recurrence after aneurysm coiling,(7, 11, 28, 29) yet there is still no definitive set of hemodynamic parameters established for clinical prediction.(30) Developing predictive parameters for coiled aneurysms is especially difficult due to the complex nature of the coil mass geometry and resulting hemodynamic variation. Coiled aneurysm hemodynamics are highly contingent on the packing density, geometry, and orientation of the final coil mass. Slight changes in coil orientation or porosity can significantly alter local fluid stresses, allow for recanalization, or even cause localized jet flow in the aneurysm, which can lead to rupture. (28, 31, 32) These phenomena are difficult predict, as it is not feasible to accurately capture coil geometry *in vivo*. The resolution required for accurately capturing the geometry of the coil mass is on the order of 10 µm, which is far beyond the resolution of typical CT scans. Additionally, imaging of coils *in vivo* is limited by the imaging artifacts caused by the presence of the coils themselves. Due to this limitation the majority of CFD studies of coiled aneurysms have relied on naïve porous media assumptions(11, 12, 33, 34) that affect hemodynamic factors in unpredictable ways.(14) While predicting aneurysm recurrence is outside of the scope of this study, Damiano et al.(35) demonstrated the potential for predicting aneurysm recurrence using CFD of virtually coiled aneurysms, representing the best attempt to relate post coiling hemodynamics to recurrence to date. That study did not compare their FEM simulations to either a ground truth (such as microtomography of *in vitro* coils) or to porous media approximations, but the results of our work above suggest a drastic improvement of the accuracy of coil resolved simulations over the porous media method, with an average error reduction of ∼58% across the hemodynamic parameters of interest.

The CFD with FEM coil geometries performed especially well in accurately capturing the average and peak velocities at the neck of the aneurysm, with average errors of 6.5% and 8.4% respectively (substantially better than the 55.0% and 36.6% average error, respectively, using the porous media approximations). This is a promising development for the ability of FEM coil geometries to improve the predictive power of CFD of coiled aneurysms, as the hemodynamics at the aneurysm neck have been implicated as especially important in predicting recurrence.(34, 35, 36)

Incorporating virtual coil geometries into CFD of coiled aneurysms using FEM is a significant improvement over the porous media method, but there is still room for improvement. As endovascular coiling is a chaotic process, even a small change in initial conditions can have a significant effect on the final coil geometry. While the general geometric agreement between the FEM and microtomography coil geometries is promising, there were still significant quantitative and qualitative differences between the coil geometries; the FEM approach captures the general shape of the coil mass, but local variations are still common. As hemodynamic parameters are sensitive to hyperlocal features of the flow, the accuracy of CFD results is not perfectly correlated with the overall accuracy of the FEM geometry. For example, patient 4 has the highest porosity correlation between the FEM and microtomography coil geometries, but also has the highest error in average WSS. This is primarily due to a small portion of the neck cross section that was not occluded in the microtomography geometry, leading to concentrated flow and high WSS in that region, as shown in Figure 2. The FEM coil geometry occluded the neck in that region and did not demonstrate the same jet effect, leading to a much lower average WSS. Even in this worst-case scenario, however, the CFD simulation using the FEM coil geometry still significantly outperformed the porous media approach.

In this work we have demonstrated the marked improvement in CFD accuracy using coil resolved simulations over the standard porous media method. Due to the large hemodynamic differences between coil resolved and porous medium CFD approaches, we recommend that future CFD based research on coiled aneurysm hemodynamics replace the homogeneous porous medium approach with a more accurate coil geometry representation when possible. We recognize that when dealing with large datasets or when computational resources are limited, conducting FEM simulations may be infeasible due to the computational cost associated with resolving the coil geometries. In this case we recommend generating localized porosity maps based on our previous work,(26) as this better approximates the porosity variation of the coil mass.

In additions to improvement in CFD simulations, there is potential for FEM simulations of endovascular coiling to aid in clinical decision-making. By examining FEM-generated coil geometries and porosity maps, clinicians may be able to select optimum coiling strategies for a specific aneurysm prior to treatment. The agreement between FEM and microtomography coil geometries in this work is promising, with an average relative error in porosity maps of only ∼8.1%, but to translate the FEM methodology for clinical use, the accuracy will need to be improved though future investigation and optimization of parameters. Precisely matching the angle and location of the catheter location during treatment is the easiest path for improving the method. In this work the catheter placement was based on angiographic imaging of the *in vivo* coil deployment, while the geometries used for comparison were from 3D-printed aneurysm models. While these models were treated in the same method and with the same coils as in actual patient treatment, it is unlikely that the catheter location and angle were identical, and this is a likely source of additional error. Furthermore, the speed of the deployment has an impact on final coil geometry.(37) In this study we did not have a record of precise deployment speed, for future study the deployment speed should match the clinical deployment speed as closely as possible. Finally, the precise effective material properties of endovascular coils are unknown, and an experimental investigation of these properties would help improve the FEM methodology.

## CONCLUSIONS

We have demonstrated the improvement in CFD simulation accuracy using FEM techniques to represent endovascular aneurysm coiling. Coil geometries generated with FEM agree well with microtomographic scans of coil geometries in 3D printed models, with an average error in porosity maps of ∼6.3%. Using FEM-generated coil geometries in CFD simulations improves the accuracy of hemodynamic calculations when compared with the standard porous media approach, with an average reduction of error in hemodynamic parameters of interest of ∼58%.

## Data Availability

All data produced in the present study are available upon reasonable request to the corresponding author

## ACKNOWLEDGMENTS

This work was supported by NIH/NINDS 1R01NS105692; an unrestricted educational grant from Stryker, which had no influence on the study design or results; and the generous support of the Catchot family.

